# Glymphatic defect in isolated REM sleep behavior disorder is associated with phenoconversion to Parkinson’s disease

**DOI:** 10.1101/2025.01.14.25320259

**Authors:** Violette Ayral, Alexandre Pastor-Bernier, Véronique Daneault, Christina Tremblay, Marie Filiatrault, Celine Haddad, Jean-François Gagnon, Ronald Postuma, Petr Dusek, Stanislav Marecek, Zsoka Varga, Johannes Klein, Michele T. Hu, Stéphane Lehéricy, Isabelle Arnulf, Marie Vidailhet, Jean-Christophe Corvol, ICEBERG Study Group, Shady Rahayel, Quebec Parkinson Network

**Affiliations:** Centre for Advanced Research in Sleep Medicine, Hôpital du Sacré-Cœur de Montréal, CIUSSS du Nord-de-l’Île-de-Montréal, Montreal H4J 1C5, Canada; Department of Neuroscience, University of Montreal, Montreal H3T 1J4, Canada; Department of Psychology, University of Montreal, Montreal H2V 2S9, Canada; Department of Psychology, University of Quebec in Montreal, Montreal H2X 3P2, Canada; Research Centre, Institut universitaire de gériatrie de Montréal, Montreal H3W 1W5, Canada; The Neuro (Montreal Neurological Institute-Hospital), McGill University, Montreal H3A 2B4, Canada; Department of Neurology, Montreal General Hospital, Montreal H3G 1A4, Canada; Department of Neurology and Centre of Clinical Neurosciences, First Faculty of Medicine, Charles University and General University Hospital, Prague, Czechia; Oxford Parkinson’s Disease Centre and Division of Neurology, Nuffield Department of Clinical Neurosciences, University of Oxford, Oxford, UK; Sorbonne Université, Institut du Cerveau – Paris Brain Institute – ICM, Assistance Publique Hôpitaux de Paris, INSERM, CNRS, Paris 75013, France; Department of Medicine, University of Montreal, Montreal H3T 1J4, Canada

**Keywords:** REM sleep behavior disorder, Parkinson’s disease, dementia with Lewy bodies, glymphatic index, diffusion MRI, prognosis, biomarker

## Abstract

Isolated rapid eye movement (REM) sleep behavior disorder (iRBD) is characterized by the loss of muscle atonia and abnormal, often violent, movements and vocalizations during REM sleep. It is the strongest prodromal marker for progression to synucleinopathies such as dementia with Lewy bodies (DLB) and Parkinson’s disease (PD). iRBD individuals already show brain changes consistent with manifest synucleinopathies, but their mechanisms remain poorly understood. The glymphatic system is involved in the brain clearance of waste products and its defect has been associated with a higher likelihood of pathological burden and neurodegeneration. The presence of glymphatic system dysfunction revealed by neuroimaging in iRBD and its association with disease progression remain uninvestigated, including its potential to predict conversion toward distinct trajectories of synucleinopathies.

We analyzed diffusion-weighted imaging data from a large international multicentric cohort of polysomnography-confirmed iRBD individuals and healthy controls. We used diffusion tensor imaging along the perivascular space to assess glymphatic function and derived a glymphatic index based on the diffusivity occurring within masks placed on associative and projection fibers adjacent to the lateral ventricles. The index was compared between groups and correlated with motor and cognitive features. Cox regression models assessed the relationship between glymphatic index and the likelihood of conversion to PD, DLB or remaining disease-free.

Our analyses included 250 polysomnography-confirmed iRBD participants (mean age 66.5 ± 6.8 years; 87% men) and 178 controls (65.7 ± 6.8 years; 81% men). There was a significantly reduced glymphatic index in iRBD individuals compared to controls. Among the 224 iRBD individuals followed longitudinally (6.1 years, 1-16 years), 65 developed a neurodegenerative disease. iRBD converters exhibited lower glymphatic index compared to non-converters. Lower glymphatic index was associated with a higher risk of phenoconversion towards PD over time compared to remaining disease-free (hazard ratio = 2.43, 95% CI = 1.13-5.25, P = 0.012).

This study reports glymphatic dysfunction in iRBD and its potential for predicting conversion to PD, underscoring its utility in identifying at-risk iRBD individuals.

## Introduction

Isolated/idiopathic rapid eye movement (REM) sleep behavior disorder (iRBD) is a parasomnia characterized by the loss of normal muscle atonia during rapid eye movement and the onset of abnormal and often violent movements and vocalizations.^1^ Approximately 90% of individuals with iRBD will develop either DLB, PD or multiple system atrophy after 15 years.^2^ Although several studies show that iRBD individuals already display brain changes reminiscent of overt synucleinopathies,^3–7^ little is known about the neurodegenerative mechanisms underlying the progression from iRBD to overt synucleinopathies.

Synucleinopathies are characterized by abnormal accumulation of alpha-synuclein, which aggregates and disrupts cellular function.^8^ This build-up of proteins has been linked to impaired brain clearance mechanisms normally helping remove toxic waste products.^9^ One hypothesized clearance pathway is the glymphatic system, a brain-wide network facilitating waste removal.^10^ In this system, primarily active during non-rapid eye movement sleep,^11,12^ cerebrospinal fluid enters the brain through periarterial routes and is cleared along perivenous pathways.^13,14^ The glymphatic system allows cerebrospinal fluid to flow into perivascular spaces around arteries through diffusion and pulsatile flow, where it mixes with interstitial fluid surrounding neurons, carrying away waste, ions, neurotransmitters, and molecules like neuropeptides that influence neuronal function. Central to this process is aquaporin-4, a protein located in astrocytes, supporting glial cells in the brain. Astrocytes extend their vascular endfeet to blood vessels, where aquaporin-4 facilitates the movement of cerebrospinal fluid between perivascular spaces and interstitial fluid, enabling efficient waste clearance. The waste-laden fluids are ultimately transported out of the brain along perivascular spaces around veins and drained into meningeal and deep cervical lymphatic systems outside the brain, supporting brain health.

Defects in the glymphatic system have been associated with the development and progression of several neurodegenerative diseases,^15,16^ including PD, Alzheimer’s disease, and amyotrophic lateral sclerosis.^11,17–19^ In these conditions, impaired glymphatic function may lead to the accumulation of harmful proteins and waste products in the brain,^20^ suggesting impaired clearance mechanisms as a contributing factor of pathology and neurodegeneration. Advanced diffusion MRI techniques, specifically diffusion tensor imaging along the perivascular space (DTI-ALPS), provide a non-invasive approach to assess interstitial fluid dynamics by measuring water diffusivity in the periventricular space.^17^ Studies using this proxy have identified glymphatic system alterations in PD and DLB, correlating with motor and cognitive impairments in PD.^18,21,22,23^ Interestingly, a unilateral onset of glymphatic index impairment has been reported in PD, being found on the left side in early de novo cases and then later involving the right side as disease progresses.^18^ However, it remains unclear whether these changes are exclusive to the overt phase of synucleinopathies or whether they may emerge in the prodromal stage.

In iRBD, single-site studies have suggested the presence of a reduced DTI-ALPS index compared to controls.^24,25,26^ However, two of these studies were conducted on limited samples of polysomnography-confirmed iRBD patients (18 and 20 patients),^25,26^ and neither explored the associations between the DTI-ALPS index and cognitive or motor dysfunction. Moreover, while Bae and colleagues proposed that glymphatic dysfunction might influence phenoconversion from iRBD to synucleinopathies,^26^ it remains unknown whether glymphatic index alterations predict differential phenoconversion towards PD or DLB in iRBD. Addressing this gap is important, as predictive biomarkers for specific neurodegenerative trajectories in iRBD are needed to monitor disease progression and to create homogeneous groups with similar phenotypes for clinical trials.^27^

In this study, we leveraged a large, multicentric dataset of diffusion-weighted brain MRI scans from 276 individuals with PSG-confirmed iRBD and 195 controls, followed longitudinally in five sites worldwide. We applied advanced diffusion MRI processing to extract the DTI-ALPS index from each participant and compared indices between iRBD individuals and controls. Using data from the latest clinical follow-up (mean follow-up duration of 6.1 years, ranging between 1-16 years), we investigated whether the DTI-ALPS index differentially predicted the development of PD versus DLB or remaining disease-free. We hypothesized that the DTI-ALPS index would be significantly lower in iRBD individuals compared to controls and that glymphatic dysfunction would associate with phenoconversion towards an overt synucleinopathy in iRBD.

## Materials and Methods

### Participants

The multicenter iRBD cohort initially included 592 participants, comprising 289 polysomnography-confirmed iRBD individuals and 303 healthy controls. After age- and sex-balancing, a total of 471 participants were included, namely 276 individuals with iRBD and 195 controls. Participants were recruited from five sites: 129 (75 patients) from the Centre for Advanced Research on Sleep Medicine at the CIUSSS-NÎM – Hôpital du Sacré-Coeur de Montréal, Montreal, Canada; 120 (73 patients) from the Oxford Discovery Cohort, Oxford, UK; 119 (74 patients) from First Faculty of Medicine at Charles University, Prague, Czechia; 76 (45 patients) from the Movement Disorders clinic (ICEBERG and ALICE Cohorts) at the Hôpital de la Pitié-Salpêtrière, Paris, France; and 27 (9 patients) from the Parkinson’s Progression Markers Initiative study (https://www.ppmi-info.org/).^28^ Recruitment was conducted independently for each site, and all iRBD patients were part of ongoing prospective longitudinal cohorts. All iRBD patients had a video-polysomnography-confirmed diagnosis according to the third edition of the International Classification of Sleep Disorders,^29^ and were free of PD, DLB, and multiple system atrophy based on established diagnostic criteria at the clinical evaluation closest in time to MRI.^30,31,32^ Each iRBD patient was followed longitudinally with annual neurological and cognitive assessments to identify the point of phenoconversion to a clinically defined overt synucleinopathy. All participants underwent the Montreal Cognitive Assessment (MoCA) to assess global cognition,^33^ and the motor scale of the Movement Disorder Society – Unified Parkinson’s Disease Rating Scale (MDS-UPDRS-III) to evaluate the severity of parkinsonian motor features.^34^ All participants were enrolled under research protocols approved by local ethics committees and provided written informed consent. This multicentric project received additional approval from the Research Ethics Board of the CIUSSS-NÎM – Hôpital du Sacré-Coeur de Montréal and the McGill University Health Center.

### Data acquisition

Diffusion-weighted MRI scans were acquired from the Montreal cohort were acquired using a 3T Siemens TIM Trio scanner with a 12-channel head coil and echo-planar sequence with the following acquisition parameters: b-values of 0 and 700 s/mm^2^; gradient directions of 63 and 64; isotropic voxel size of 2 mm; TR ranging from 8.6 to 12.7 ms; TE ranging from 0.083 to 0.1 ms; or using a 3T Siemens PRISMA scanner with a 32-channel head coil and echo-planar sequence: b-values of 0 and 1000 s/mm^2^; gradient directions of 30; isotropic voxel size of 2 mm; TR = 6.9 ms; TE = 64 ms. The Oxford cohort was scanned using a 3T Siemens Trio scanner with a 12-channel head coil and echo-planar sequence: b-values of 0 and 1000 s/mm^2^; gradient directions of 60; isotropic voxel size of 2 mm; TR = 9.3 ms; TE = 94 ms. The Prague cohort was scanned using a 3T Siemens Skyra with a 32-channel head coil and echo-planar sequence: b-values of 0 and 1000 s/mm^2^; gradient directions of 29 or 30; isotropic voxel size of 2 mm; TR = 10.5 ms; TE = 93 ms. The Paris cohort was scanned using a 3T Siemens TIM Trio scanner with a 12-channel head coil and echo-planar imaging: b-values of 0 and 700 s/mm^2^; gradient directions of 21 directions; isotropic voxel size of 1.72 mm; TR = 14 ms; TE = 101 ms; or using a 3T PRISMA Fit scanner with a 64-channel head coil and echo-planar imaging: b-values of 0 and 700 s/mm^2^; gradient directions of 32 directions; isotropic voxel size of 1.7 mm; TR = 10.4 ms; TE = 59 ms.

The acquisition parameters for the T1-weighted MRI scans from the Montreal cohort were acquired with an MPRAGE sequence with the following parameters: TR = 2300 ms; TE = 2.91 ms; flip angle = 9°; and isotropic voxel size of 1 mm; or with TR = 2300 ms; TE = 2.98 ms; flip angle = 9°; and isotropic voxel size of 1 mm. The Oxford cohort used MPRAGE with TR = 2040 ms; TE = 4.7 ms; flip angle = 8°; and isotropic voxel size of 1 mm. The Prague cohort used MPRAGE sequence with: TR = 2200 ms; TE = 2.4 ms; flip angle = 8°; and isotropic voxel size of 1 mm. The Paris cohort used MPRAGE sequence with: TR = 2300 ms; TE = 4.18 ms; flip angle = 9°; and isotropic voxel size of 1 mm; or MP2RAGE with: TR = 5000 ms; TE = 2.98 ms; flip angles = 4° and 5°; GRAPPA = 3; and isotropic voxel size of 1 mm. The acquisition parameters for the Parkinson’s Progression Markers Initiative study have been described elsewhere.^32^

### Diffusion MRI processing

Diffusion-weighted brain MRI scans were processed using the Tractoflow-ABS automated pipeline (see Figure 1 for the imaging protocol).^35,36^ The brain extraction tool (BET) was used to remove non-brain tissue from the images, and eddy currents and head motion artefacts were corrected using the eddy tool. A brain mask was extracted from the non-diffusion-weighted (b0) image and the diffusion tensor model was fit to the corrected data with the dtifit tool. This generated fractional anisotropy (FA), mean diffusivity, and directional diffusivity maps along the left-right direction (Dxx), anterior-posterior direction (Dyy), and superior-inferior direction (Dzz). All images were visually inspected using FSLeyes and data with significant artefacts were excluded.^37^

**Figure 1.**
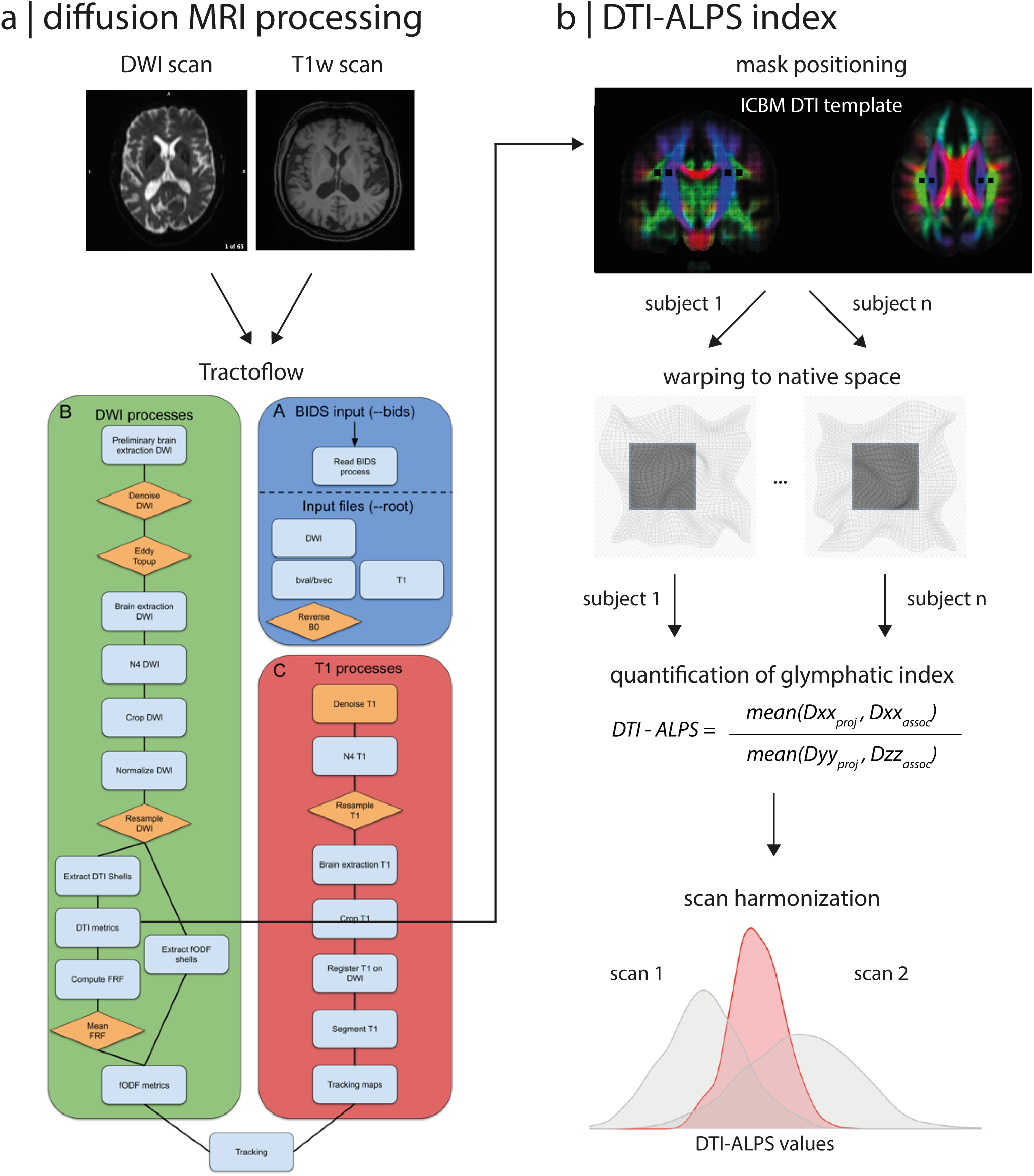
Processing steps involved in deriving the glymphatic index. **a)** Diffusion- and T1-weighted MRI scans were processed using the Tractoflow-ABS pipeline. **b)** Using the fractional anisotropy map template from the ICBM DTI-81 atlas, four masks were positioned on the associative and projection fibers at the level of the lateral ventricle body of each hemisphere and were warped to every participant’s native space. Diffusivity measurements were extracted and harmonized for scanner effects using ComBat. The DTI-ALPS index was calculated for each participant as a proxy of glymphatic function. ABS = Atlas-Based Segmentation; ALPS = glymphatic (‘along the perivascular space’) index; DTI = diffusion tensor imaging; DWI = diffusion-weighted imaging; iRBD = isolated rapid eye movement sleep behavior disorder; T1w = T1-weighted MRI scan.

For quantitative DTI-ALPS index derivation, measurements were extracted from the diffusivity maps. Using the FA map template from the ICBM DTI-81 atlas,^38,39^, we generated four 5-mm cubic masks that were positioned on associative (MNI coordinates: x = 39, y = -17, z = 29 on the left and x = -39, y = -17, z = 29 on the right) and projection fibers (MNI coordinates: x = 26, y = -17, z = 29 on the left and x = -26, y = -17, z = 29 on the right) at the level of the lateral ventricle body in each hemisphere, as done previously.^17^ These masks were registered to each participant’s native space using Advanced Normalization Tools (https://stnava.github.io/ANTs) and visual quality control was done on the RGB map to ensure accurate coverage of the regions of interest. The masks were then applied to each participant’s Dxx, Dyy, and Dzz maps from Tractoflow to extract diffusivity measurements for left and right association and projection fiber masks. For each participant, the DTI-ALPS index was calculated as follows, based on Taoka and colleagues:^17^

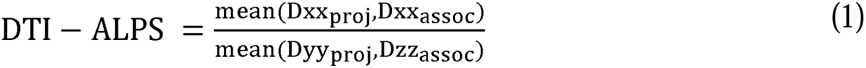

To account for the multicentric nature of this dataset and the varying acquisition parameters across different scanners, we used the ComBat algorithm.^40,41^ ComBat is a standard tool in neuroimaging studies for harmonizing data across sites and has been validated in previous studies using T1- and diffusion-weighted MRI scans.^40^

### Statistical Analysis

Statistical analyses were conducted using IBM SPSS software (version 29.0.1.0). Independent samples t-tests were used to compare age and follow-up time between iRBD and control groups, as well as to assess whether DTI-ALPS indices (left and right) were lower in iRBD individuals compared to controls. Chi-squared tests were used to assess differences in sex distribution. For non-normally distributed variables (MoCA and MDS-UPDRS-III), the Mann-Whitney U test was used. Pearson’s correlation coefficients were calculated to investigate associations between the DTI-ALPS indices (left and right) and clinical performance on the MoCA and MDS-UPDRS-III. Independent samples t-tests further assessed whether DTI-ALPS indices were decreased in iRBD that converted to an overt synucleinopathy (converters) compared to age- and sex-balanced iRBD who remained disease-free (non-converters). A binary regression was applied to examine whether the DTI-ALPS indices associated with the likelihood of differential progression trajectories, namely phenoconversion to PD or phenoconversion to DLB compared to non-converters, while controlling for age and sex. To determine the added predictive value of DTI-ALPS indices relative to traditional assessments, additional binary regression models assessed whether the DTI-ALPS indices associated with phenoconversion trajectories beyond standard clinical measures (MoCA and MDS-UPDRS-III), while controlling for age and sex. Kaplan-Meier analysis was performed the association between glymphatic index and risk of phenoconversion over time. Time 0 represented the time at which the brain MRI was performed, and the distance between the MRI and the date of the clinical evaluation where disease onset was confirmed (or the latest clinical evaluation for patients still disease-free) was calculated for each patient. For assessing risk of phenoconversion, patients were split into high and low left ALPS index based on the median ALPS index value for the iRBD group. The primary analysis involved assessing whether there was a difference in survival between high and low left ALPS index groups of iRBD patients, with the log-rank test used to determine statistical significance. Additional analyses were conducted to assess differences in the risk over time of PD versus disease-free and DLB versus disease-free trajectories. Cox proportional hazards models were applied to examine the adjusted impact of the DTI-ALPS indices on phenoconversion to PD or DLB over time, while controlling for age and sex as covariates. Likelihood ratio tests were used to assess model significance. DTI-ALPS indices were z-scored before being entered in the regression models to ease interpretability of a one-unit change in the odds and hazard ratios.

## Results

### Participants

After quality control, 43 participants (9.3%, 26 iRBD individuals) were excluded due to issues such as diffusion-weighted imaging processing failures, mask misregistration over fibers, and extreme values, yielding a final sample for analysis of 250 iRBD individuals and 178 controls. There were no significant differences between the remaining iRBD and control groups in terms of age at MRI acquisition (P = 0.26) or sex distribution (P = 0.10). iRBD patients showed significantly lower MoCA scores (iRBD: 25.4 ± 3.0 versus controls: 26.6 ± 2.3, P < 0.001) and higher MDS-UPDRS-III scores (iRBD: 6.3 ± 5.6 versus controls: 3.8 ± 6.0, P < 0.001) compared to controls (Table 1).

**Table 1.** Demographic, clinical, and DTI-ALPS variables of iRBD patients and controls.

| Variables | iRBD (n=250) | Controls (n=178) | P-value |
| --- | --- | --- | --- |
| Age at MRI, years | 66.5 ± 6.8 | 65.7 ± 6.8 | 0.25<sup>a</sup> |
| Sex, N (% men) | 217 (87%) | 144 (81%) | 0.10<sup>b</sup> |
| MoCA | 25.4 ± 3.0 | 26.6 ± 2.3 | **<0.001<sup>c</sup>** |
| MDS-UPDRS-III | 6.3 ± 5.6 | 3.8 ± 6.0 | **<0.001<sup>c</sup>** |
| DTI-ALPS index,<br>left | 1.288 ± 0.198 | 1.322 ± 0.203 | **0.043<sup>a</sup>** |
| DTI-ALPS index,<br>right | 1.264 ± 0.181 | 1.286 ± 0.185 | 0.10<sup>a</sup> |

Values are presented as mean ± standard deviation. Bold values represent significant differences.
<sup>a</sup> Student's independent two-sample t-test.
<sup>b</sup> Chi-squared test.
<sup>c</sup> Mann-Whitney U test.
ALPS = glymphatic ('along the perivascular space') index; DTI = diffusion tensor imaging; iRBD = isolated rapid eye movement sleep behavior disorder; MDS = Movement Disorder Society; MoCA = Montreal Cognitive Assessment; UPDRS-III = motor part of the Unified Parkinson's Disease Rating Scale.

### The DTI-ALPS index is decreased in iRBD

We first investigated whether the DTI-ALPS index differed between iRBD patients and controls. Independent samples t-tests revealed a significantly lower left ALPS index in iRBD patients compared to controls, with iRBD patients having a DTI-ALPS index of 1.29 ± 0.20 compared to 1.32 ± 0.20 in controls (mean difference ± standard error: -0.034 ± 0.20, P = 0.043, Cohen’s d = 0.17) (Table 1 and Figure 2). No significant differences were found for the right DTI-ALPS index between iRBD patients and controls (P = 0.10) (Table 1 and Figure 2). These findings suggest that the glymphatic system is impaired in iRBD patients compared to controls.

**Figure 2.**
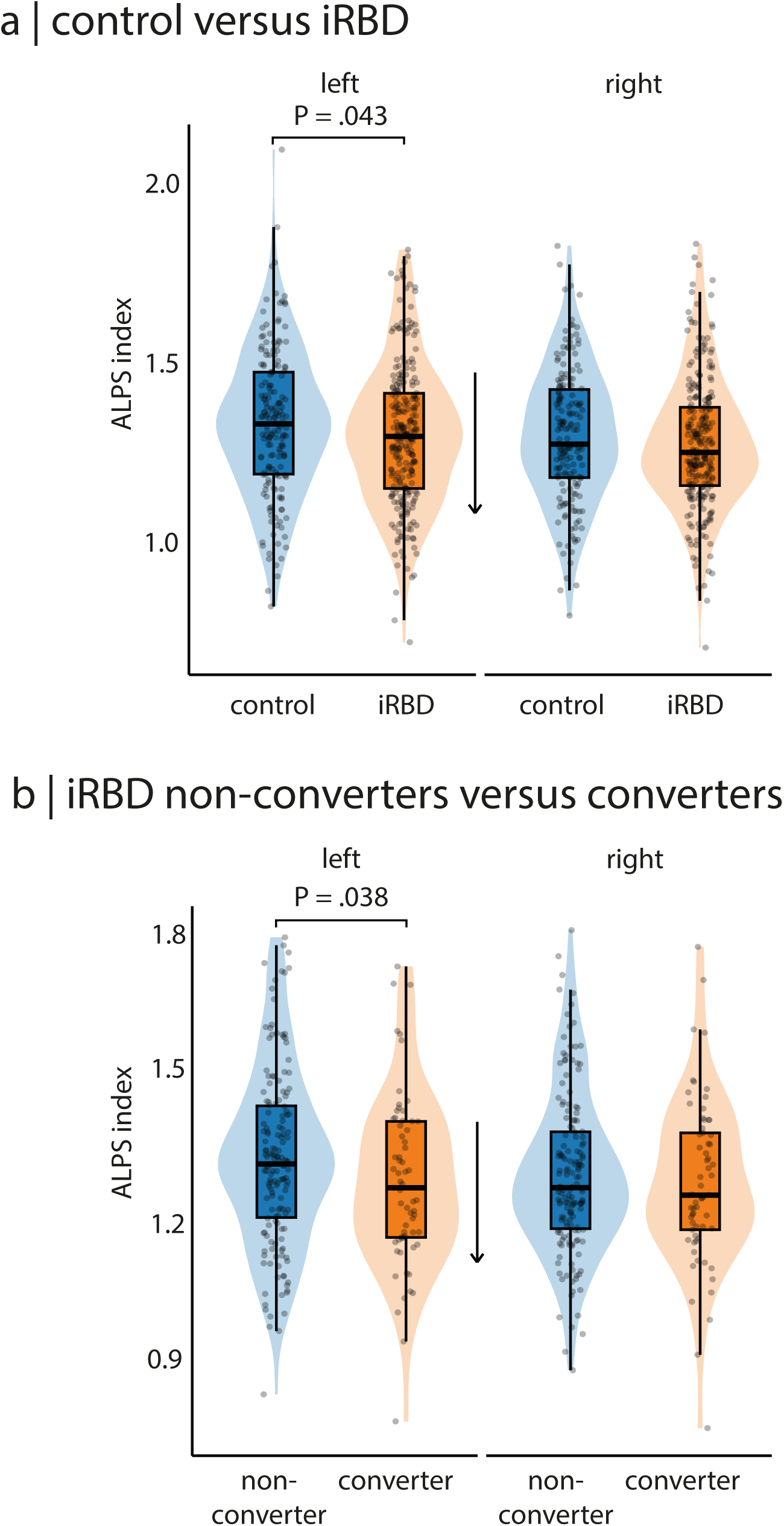
Comparison of DTI-ALPS index between iRBD individuals and controls. **a)** Violin plots showing the distribution of DTI-ALPS indices (left and right) in healthy controls (blue) and iRBD individuals (orange). The left DTI-ALPS index is significantly lower in iRBD patients compared to controls. **b)** Violin plots showing the distribution of DTI-ALPS indices (left and right) in non-converted iRBD (blue) and phenoconverted iRBD individuals (orange). The left DTI-ALPS index is significantly lower in phenoconverted iRBD patients compared to non-converted iRBD The plot includes boxplots within each violin, displaying the median (central line) and interquartile range) of DTI-ALPS indices. ALPS = glymphatic (‘along the perivascular space’) index; DTI = diffusion tensor imaging; iRBD = isolated rapid eye movement sleep behavior disorder.

### Lower DTI-ALPS index is associated with lower cognitive function

Next, we investigated whether the DTI-ALPS index was associated with clinical measures, namely global cognition (MoCA) and severity of parkinsonian motor features (MDS-UPDRS-III). In iRBD, no correlations were found between MoCA scores and the left (r = 0.07, P = 0.26) and right (r = 0.07, P = 0.31) DTI-ALPS indices. Similarly, in controls, no correlations were found between MoCA scores and the left (r = 0.14, P = 0.09) and right (r = 0.12, P= 0.15) DTI-ALPS indices. When considering all participants, a significant positive correlation was found between MoCA scores and the left (r = 0.116, P = 0.024) DTI-ALPS index (Figure 3), but did not remain significant when correcting for age and sex (r = 0.07, P = 0.16). No significant correlation was observed between MoCA scores and the right DTI-ALPS index (r = 0.096, P = 0.060). There were also no significant correlations between the MDS-UPDRS-III scores and the left and right DTI-ALPS indices in the whole cohort (P > 0.30), and when looking at controls (P > 0.30) and iRBD groups independently (P > 0.14).

**Figure 3.**
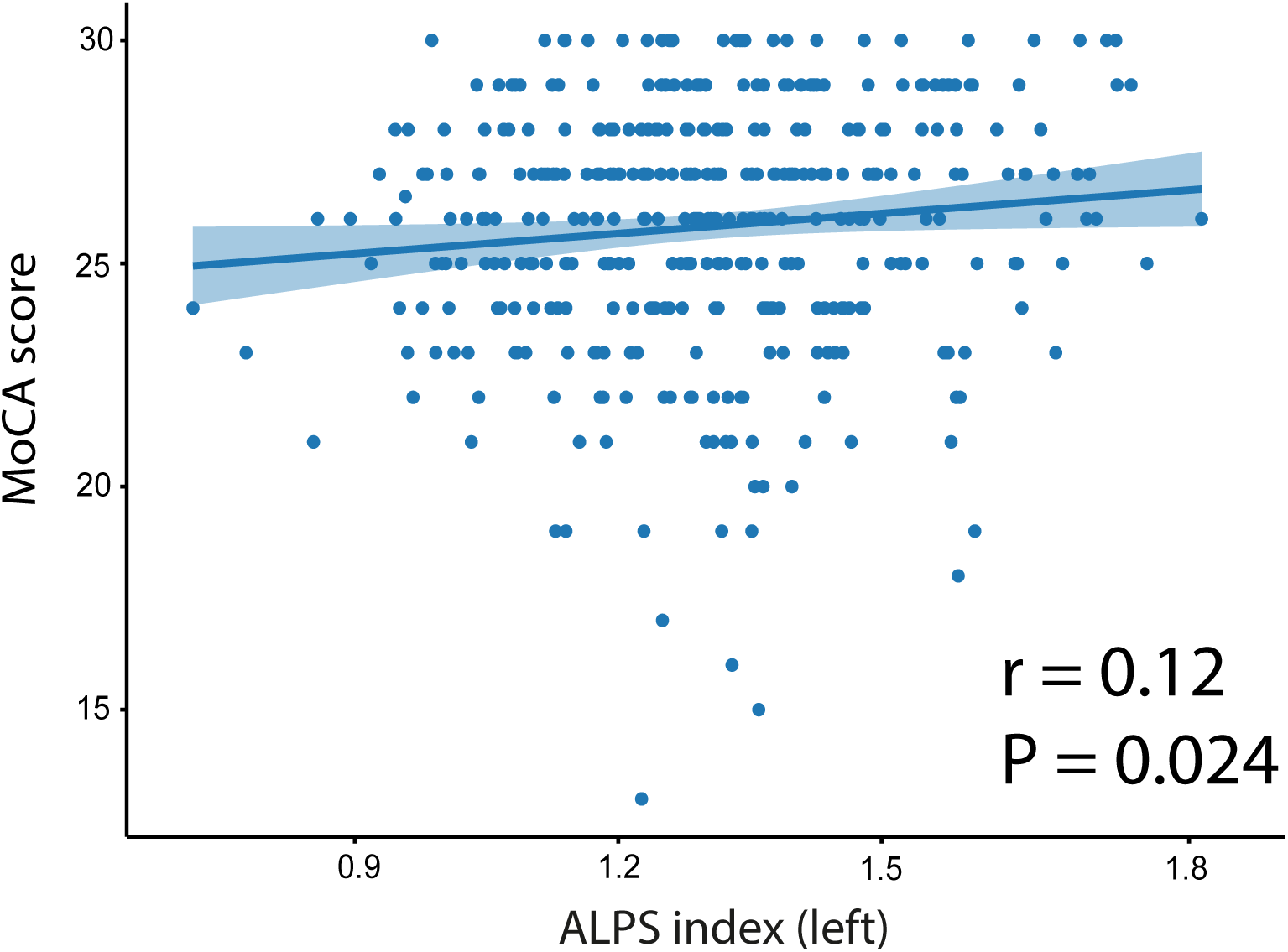
Correlation between DTI-ALPS index and MoCA scores. Scatter plot illustrating the relationship between the left DTI-ALPS index and global cognitive function, as measured by the MoCA. ALPS = glymphatic (‘along the perivascular space’) index; DTI = diffusion tensor imaging; MoCA = Montreal Cognitive Assessment.

### Lower DTI-ALPS index associates with phenoconversion in iRBD

Of the 250 iRBD patients, 224 (90%) were followed longitudinally with annual neurological and cognitive assessments over a mean duration of 6.1 years (range: 1-16 years). At the latest clinical follow-up, 65 individuals (29%) had developed an overt synucleinopathy (converters), while 159 individuals (71%) remained disease-free (non-converters). Converters and non-converters did not significantly differ in age at MRI (P = 0.81), sex distribution (P = 0.80), and MDS-UPDRS-III scores (P = 0.24). However, converters had significantly lower MoCA scores (mean difference ± standard error: 24.9 ± 2.9 versus 25.9 ± 2.8, P = 0.011) (Table 2). When comparing DTI-ALPS indices between iRBD converters and non-converters, an independent samples t-test revealed that converters had a significantly lower left DTI-ALPS index compared to non-converters (1.25 ± 0.19 versus 1.30 ± 0.20, P = 0.038, Cohen’s d = 0.26) (Figure 2 and Table 2). No significant differences were found for the right DTI-ALPS index (P = 0.24) (Figure 2 and Table 2).

**Table 2.** Demographic, clinical, and DTI-ALPS variables of iRBD converters and non-converters.

| Variables | iRBD non-converters<br>(n=159) | iRBD converters<br>(n=65) | P-value |
| --- | --- | --- | --- |
| Age at MRI, years | 66.2 ± 7.2 | 66.4 ± 6.2 | 0.81<sup>a</sup> |
| Sex, N (% men) | 140 (88%) | 58 (89%) | 0.80<sup>b</sup> |
| MoCA | 25.9 ± 2.8 | 24.9 ± 2.9 | **0.011**<sup>c</sup> |
| MDS-UPDRS-III | 6.2 ± 5.9 | 6.8 ± 5.7 | 0.24<sup>c</sup> |
| DTI-ALPS index, left | 1.304 ± 0.198 | 1.254 ± 0.193 | **0.038**<sup>a</sup> |
| DTI-ALPS index, right | 1.269 ± 0.181 | 1.250 ± 0.186 | 0.24<sup>a</sup> |

Values are presented as mean ± standard deviation. Bold values represent significant differences.
<sup>a</sup> Student's independent two-sample t-test.
<sup>b</sup> Chi-squared test.
<sup>c</sup> Mann-Whitney U test.
ALPS = glymphatic ('along the perivascular space') index; DTI = diffusion tensor imaging; iRBD = isolated rapid eye movement sleep behavior disorder; MDS = Movement Disorder Society; MoCA = Montreal Cognitive Assessment; UPDRS-III = motor part of the Unified Parkinson's Disease Rating Scale.

### Lower DTI-ALPS index specifically associates with PD

Among the 65 iRBD converters, 42 individuals (65%) had developed PD, 18 (28%) DLB, 4 (6%) multiple system atrophy, and 1 (2%) AD at the latest clinical follow-up. To evaluate whether DTI-ALPS indices associated with different trajectories in iRBD, specifically phenoconversion to PD or DLB as opposed to remaining disease-free, we conducted logistic regressions, considering separately iRBD converters to PD or DLB. Both groups did not differ in age (PD: 66.6 ± 6.1 versus DLB: 67.6 ± 5.7, P = 0.58), sex distribution (93% men in PD versus 83% men in DLB, P = 0.26), and MDS-UPDRS-III scores (PD: 6.6 ± 5.2 versus DLB: 8.6 ± 7.0, P = 0.24). As expected, DLB converters had significantly lower MoCA scores compared to PD converters (23.1 ± 2.7 versus 25.7 ± 2.7, P < 0.001). In the first model, which included the left DTI-ALPS index, age, and sex as predictors, the left DTI-ALPS index was identified as a significant predictor for PD conversion compared to remaining disease-free (B = -0.42, standard error = 0.20, P = 0.029) (Table 3). The odds ratio was 0.65 (95% CI: 0.45, 0.96) for the left DTI-ALPS index, indicating that each one-standard deviation increase in the index (i.e., healthier glymphatic function) reduced the odds of converting to PD by 35% compared to staying disease-free. In contrast, the left DTI-ALPS index did not significantly associate with DLB conversion (P = 0.90) (Table 3). The right DTI-ALPS index was not a significant predictor for conversion to either PD (P = 0.33) or DLB (P = 0.65). Similar results were found when assessing conversion to PD or conversion to DLB compared to any other longitudinal trajectory in iRBD (i.e., disease-free or other) (Table S1).

**Table 3.** Models of the association between ALPS index and trajectories in iRBD.

| Model Term | Conversion to PD<sup>a</sup> |  |  | Conversion to DLB<sup>a</sup> |  |  |
| --- | --- | --- | --- | --- | --- | --- |
|  | Coefficient (SE) | P-value | Odds ratio (95% CI) | Coefficient (SE) | P-value | Odds ratio (95% CI) |
| *Model 1: ALPS* |  |  |  |  |  |  |
| DTI-ALPS index, left | -0.424 (0.20) | **0.029** | 0.654 (0.447-0.958) | 0.03 (0.26) | 0.90 | 1.03 (0.62-1.71) |
| Age | -0.003 (0.03) | 0.92 | 1.00 (0.95-1.05) | 0.03 (0.04) | 0.42 | 1.03 (0.96-1.11) |
| Sex | -0.27 (0.67) | 0.68 | 0.76 (0.21-2.80) | 0.37 (0.70) | 0.59 | 1.45 (0.37-5.70) |
| Constant | -1.19 (1.77) | 0.50 | 0.30 | -4.22 (2.51) | 0.09 | 0.02 |
| *Model 2: MoCA* |  |  |  |  |  |  |
| DTI-ALPS index, left | -0.389 (0.19) | **0.045** | 0.678 (0.46-0.99) | -0.004 (0.28) | 0.99 | 1.00 (0.58-1.72) |
| Age | 0.003 (0.03) | 0.92 | 1.00 (0.95-1.06) | 0.01 (0.04) | 0.80 | 1.01 (0.93-1.09) |
| Sex | -0.13 (0.68) | 0.84 | 0.88 (0.23-3.30) | 0.45 (0.77) | 0.56 | 1.56 (0.35-7.03) |
| MoCA | -0.003 (0.07) | 0.97 | 1.00 (0.88-1.14) | -0.28 (0.09) | **0.001** | 0.75 (0.64-0.89) |
| Constant | -1.45 (2.71) | 0.59 | 0.24 | 4.10 (3.62) | 0.26 | 60.26 |
| *Model 3: MDS-UPDRS-III* |  |  |  |  |  |  |
| DTI-ALPS index, left | -0.41 (0.20) | **0.035** | 0.66 (0.45-0.97) | -0.03 (0.27) | 0.91 | 0.97 (0.58-1.63) |
| Age | -0.01 (0.03) | 0.67 | 0.98 (0.94-1.04) | 0.024 (0.04) | 0.56 | 1.02 (0.95-1.11) |
| Sex | -0.19 (0.68) | 0.78 | 0.83 (0.22-3.13) | 0.48 (0.73) | 0.51 | 1.61 (0.39-6.71) |
| MDS-UPDRS-III | 0.01 (0.03) | 0.73 | 1.01 (0.95-1.08) | 0.05 (0.04) | 0.19 | 1.05 (0.98-1.14) |
| Constant | -0.58 (1.82) | 0.75 | 0.56 | -4.16 (2.70) | 0.12 | 0.02 |

Bold values represent significant predictors.
<sup>a</sup> Tested against staying disease-free in longitudinally followed iRBD
ALPS = glymphatic ('along the perivascular space') index; CI = confidence interval; DLB = dementia with Lewy bodies; DTI = diffusion tensor imaging; iRBD = isolated rapid eye movement sleep behavior disorder; MDS = Movement Disorder Society; MoCA = Montreal Cognitive Assessment; PD = Parkinson's disease; SE = standard error; UPDRS-III = motor part of the Unified Parkinson's Disease Rating Scale.

When cognitive scores (MoCA) were added to the model to assess whether glymphatic index provided more information than typical assessments, the left DTI-ALPS index remained a significant predictor of phenoconversion to PD (B = -0.39, standard error = 0.19, P = 0.045, odds ratio = 0.68, 95% CI = 0.46-0.99) (Table 3). With this additional variable, a one-standard deviation increase of the left DTI-ALPS index reduced the odds of converting to PD by 32%, translating to 1.5 times higher odds of PD conversion. The left DTI-ALPS index still did not predict conversion to DLB (P = 0.99) (Table 3). In a third model including motor scores (MDS-UPDRS-III) instead, the left DTI-ALPS index also remained a significant predictor of the development of PD (B = -0.41, standard error = 0.20, P = 0.035, odds ratio = 0.66, 95% CI = 0.45, 0.97) (Table 3). A one-standard deviation increase in the left DTI-ALPS index showed a reduced odds of converting to PD by 34%, also translating to 1.5 times lower odds of conversion to PD. Again, the left DTI-ALPS index did not predict DLB conversion (P = 0.91) (Table 3).

To assess the association between glymphatic index and risk of phenoconversion over time, Kaplan-Meier survival analysis was performed. Patients were categorized into high and low left ALPS index groups based on the median ALPS index value in iRBD. Survival curves revealed a significant difference in the proportion of iRBD patients who remained disease-free between the high and low ALPS index groups, with those having a low index demonstrating a higher risk of phenoconverting over time compared to those with high index (P = 0.034) (Figure 4). A Cox proportional hazards regression model further indicated that lower ALPS index was associated with an increased risk of phenoconverting over time (HR = 1.93, 95% CI: 1.05-3.57, P = 0.035), independently from age and sex (Table S2).

**Figure 4.**
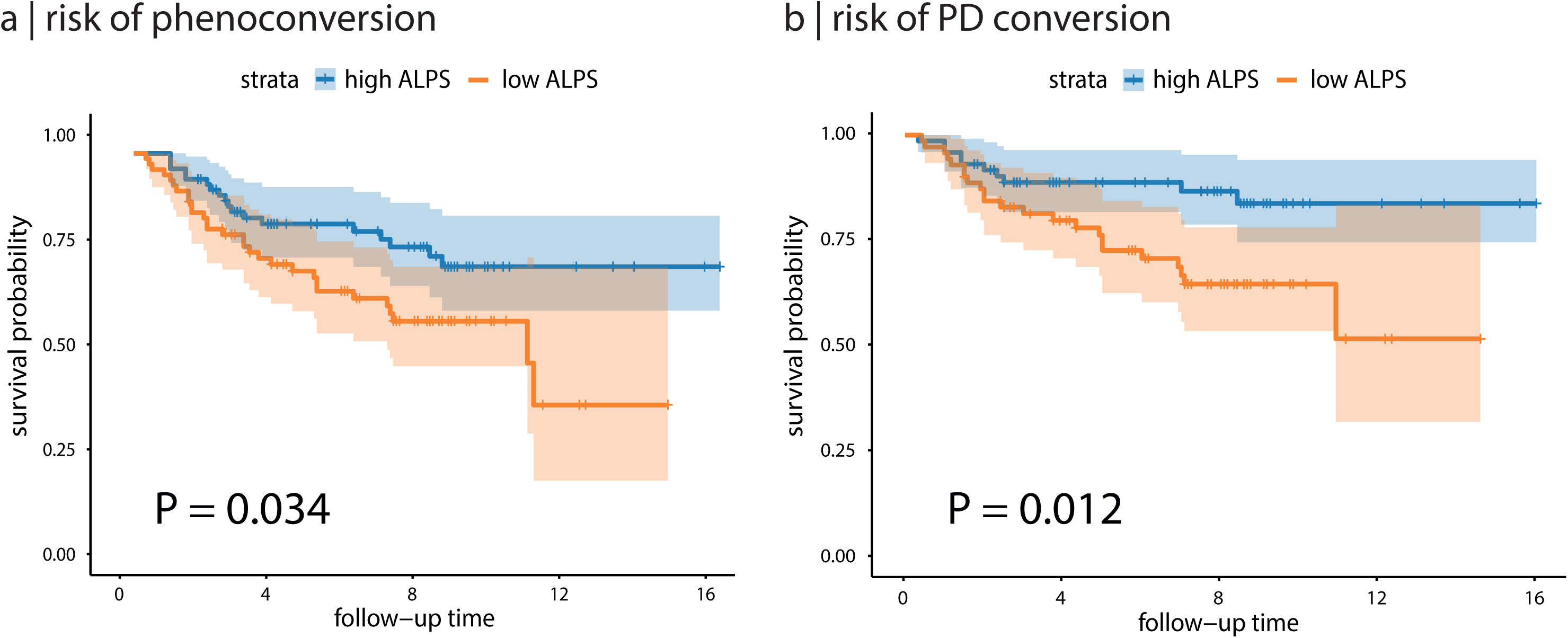
Kaplan-Meier survival curves of conversion based on ALPS index. **a)** Kaplan-Meier survival curves illustrating the relationship between low and high ALPS index (based on the median) and conversion risk in iRBD. A lower left ALPS Z-score is associated with a higher risk of conversion. **b)** Kaplan-Meier survival curves illustrating the relationship between low and high ALPS index (based on the median) and conversion risk to PD in iRBD. A lower left ALPS Z-score is associated with a higher risk of conversion to PD. Shaded areas represent the 95% confidence intervals. ALPS = glymphatic (‘along the perivascular space’) index; PD = Parkinson’s disease.

When assessing specific phenoconversion trajectories, Kaplan-Meier analysis revealed that a lower left ALPS index in iRBD was significantly associated with a higher risk of phenoconversion towards PD over time compared to remaining disease-free (P = 0.012) (Figure 4). This was independent from the effects of age and sex based on the Cox proportional hazards model (HR = 2.43, 95% CI: 1.13-5.25, P = 0.023) (Table S3). In contrast, the left ALPS index did not significantly predict phenoconversion towards DLB compared to remaining disease-free in iRBD (P = 0.89) (Figure S1 and Table S4).

## Discussion

In this study, we investigated glymphatic dysfunction in a large, multicentric cohort of individuals with polysomnography-confirmed iRBD. Our findings provide evidence that glymphatic dysfunction, measured by the DTI-ALPS index, is present in this population. Furthermore, iRBD patients who phenoconverted to an overt neurodegenerative synucleinopathy during follow-up had a lower left DTI-ALPS index at baseline compared to those who remained disease-free. Regression analyses identified the left DTI-ALPS index as a significant predictor of phenoconversion, specifically towards PD, compared to staying disease-free. This effect was present independently from typical clinical assessments such as the MoCA and MDS-UPDRS-III. Taken together, these results indicate that glymphatic dysfunction in iRBD may serve as a potential predictive marker for progression to manifest synucleinopathies, specifically PD.

Previous studies reported glymphatic dysfunction using DTI-ALPS in various neurocognitive disorders. In PD, studies consistently reported a lower DTI-ALPS index in patients relative to controls, associated with both motor and cognitive impairments.^18,21,22^ Specifically, lower DTI-ALPS index was associated with lower MMSE scores and higher MDS-UPDRS-III scores in multiple studies.^18,21,22,42^ A lower DTI-ALPS index was also recently observed in DLB patients compared to controls.^23^ However, research specifically addressing glymphatic dysfunction in iRBD remains limited.^24–26^ The existing studies on iRBD were conducted on small, single-center cohorts, some relying on questionnaire-based diagnoses of iRBD, which may be prone to false positives,^43^ and others focusing solely on the bilateral DTI-ALPS index.^24,25,26^ A previous study on 18 iRBD patients reported a reduced DTI-ALPS index compared to controls.^25^ This finding was confirmed in another small cohort of 20 iRBD patients,^26^ and in a larger cohort of 119 iRBD patients that recruited participants with probable RBD based on questionnaires.^24^ In the present study, we assessed the presence of glymphatic dysfunction in iRBD by leveraging the largest dataset of diffusion MRI scans from patients with polysomnography-confirmed iRBD. After accounting for harmonization of glymphatic function values across multiple centers, our findings confirmed that iRBD patients exhibit reduced glymphatic function when using the DTI-ALPS approach, as in previous studies. However, in our case, this reduction was specifically observed in the left DTI-ALPS index. This is consistent with a study in PD that reported a decreased DTI-ALPS index starting in the left hemisphere, which subsequently progressively involves the right hemisphere as disease progresses.^21^ Observations of unilateral glymphatic dysfunction in iRBD may therefore represent the earliest changes of this effect in synucleinopathies. Future studies should investigate more closely how this glymphatic impairment in iRBD relates to the brain structural alterations affecting gray and white matter in iRBD patients.

iRBD individuals often convert 10 years or more after the onset of clinical symptoms.^44^ In this study, we found that the DTI-ALPS index was a significant predictor of conversion trajectories in iRBD, specifically towards PD. This association was present even when adding standard clinical assessments within models such as the MoCA and MDS-UPDRS-III. This highlights that changes in the DTI-ALPS index predict progression in iRBD, supporting its value as an imaging biomarker in prodromal neurodegenerative conditions. Our findings align with a previous study that explored the predictive value of the DTI-ALPS index in iRBD patients and showed that a lower index associated with disease progression, although specific disease trajectories were not identified.^26^ These findings also align with several brain imaging studies in iRBD showing the presence of cortical and subcortical atrophy and abnormal patterns of brain perfusion that can predict specific disease trajectories.^45,4^ It also aligns with specific clinical features shown to predict specific phenoconversion trajectories in iRBD.^46^ Previous studies by our group show, using in silico models,^47,48^ that neurodegenerative changes found in iRBD, namely brain atrophy, can be recreated based on the spread of alpha-synuclein proteins in the brain as a function of deafferentation and the accumulation of toxic proteins in specific regions.^48^ Although only connection strength and local gene expression were used as spreading and vulnerability factors in these simulations, our study suggests that altered glymphatic function may be an additional player in such models. In addition, our observation that a lower glymphatic index predicted conversion to PD rather than DLB is intriguing and warrants further investigation. This contrasts with previously identified neuroimaging biomarkers in iRBD, where increased tissue deformation or relative hyperperfusion of the basal forebrain were specific predictors of DLB compared to PD.^4^ Taken together, these results support the idea that combining MRI-based differential markers, namely glymphatic index and brain atrophy, may allow for more specific prediction of disease trajectories, potentially improving patient stratification in clinical trials and prognosis in neurodegenerative diseases.

Some limitations should be acknowledged. While we used the largest cohort of iRBD patients available to date, only a subset of patients had yet phenoconverted. Future studies should continue to monitor these patients longitudinally to capture when they convert, and whether they convert to PD or DLB. Additionally, the low number of patients phenoconverting to other phenotypes, such as multiple system atrophy, precluded their inclusion in the current analysis of specific trajectories. There are also inherent limitations in the DTI-ALPS methodology itself. The DTI-ALPS index specifically measures diffusion in the periventricular area, which may not be sensitive enough to detect glymphatic system dysfunction across the entire brain.^24,49^ Moreover, the DTI-ALPS index reflects water diffusivity, an indirect measure of glymphatic activity, and its interpretation should be approached with caution.^50^ Nevertheless, despite this limitation, the DTI-ALPS method has shown potential in associating with clinical features in numerous neurodegenerative diseases. Indeed, even if it primarily reflects water diffusivity, we show that it still provides valuable insights into fluid dynamics that can predict specific trajectories of disease progression in iRBD. Another limitation is that, in this multicentric cohort, the only common clinical assessments across cohorts were the MoCA and MDS-UPDRS-III, which are relatively broad measures. While these assessments provide insight into global cognitive function and parkinsonian motor features, they may not capture the full spectrum of disease-related changes. Future studies should incorporate more fine-grained clinical assessments across cohorts to better understand the specific cognitive domains and motor features affected.

In summary, we show the presence of glymphatic dysfunction in a large multicentric cohort of patients with polysomnography-confirmed iRBD. We found that the DTI-ALPS index was significantly lower in iRBD patients compared to controls and was a predictor of conversion to PD. These findings suggest that glymphatic dysfunction occurs early in the PD progression continuum associated to iRBD and may play a role in disease progression.

## Acknowledgements

There are no acknowledgments for this paper.

## Data availability statement

The data used in this study were obtained from multiple collaborating centers, each of which retains ownership of their respective datasets. The principal investigator had authorized access to all data necessary for the analyses performed in this study. However, the accessibility and sharing of data are subject to the local policies and restriction criteria of each center involved. As such, data availability is restricted, and requests for access should be directed to the respective institutions, pending their specific data access and sharing guidelines.

## Appendix 1

List of the contributors involved in the ICEBERG Study Group:

Steering committee: Marie Vidailhet, MD, PhD, (Pitié-Salpêtrière Hospital, Paris, principal investigator of ICEBERG), Jean-Christophe Corvol, MD, PhD (Pitié-Salpêtrière Hospital, Paris, scientific lead), Isabelle Arnulf, MD, PhD (Pitié-Salpêtrière Hospital, Paris, member of the steering committee), Stéphane Lehericy, MD, PhD (Pitié-Salpêtrière Hospital, Paris, member of the steering committee);

Clinical data: Marie Vidailhet, MD, PhD, (Pitié-Salpêtrière Hospital, Paris, coordination), Graziella Mangone, MD, PhD (Pitié-Salpêtrière Hospital, Paris, co-coordination), Jean-Christophe Corvol, MD, PhD (Pitié-Salpêtrière Hospital, Paris), Isabelle Arnulf, MD, PhD (Pitié-Salpêtrière Hospital, Paris), Sara Sambin, MD (Pitié-Salpêtrière Hospital, Paris), Jonas Ihle, MD (Pitié-Salpêtrière Hospital, Paris), Caroline Weill, MD, (Pitié-Salpêtrière Hospital, Paris), David Grabli, MD, PhD (Pitié-Salpêtrière Hospital, Paris); Florence Cormier-Dequaire, MD (Pitié-Salpêtrière Hospital, Paris); Louise Laure Mariani, MD, PhD (Pitié-Salpêtrière Hospital, Paris), Bertrand Degos, MD, PhD (Avicenne Hospital, Bobigny); Neuropsychological data: Richard Levy, MD (Pitié-Salpêtrière Hospital, Paris, coordination), Fanny Pineau, MS (Pitié-Salpêtrière Hospital, Paris, neuropsychologist), Julie Socha, MS (Pitié-Salpêtrière Hospital, Paris, neuropsychologist), Eve Benchetrit, MS (La Timone Hospital, Marseille, neuropsychologist), Virginie Czernecki, MS (Pitié-Salpêtrière Hospital, Paris, neuropsychologist), Marie-Alexandrine, MS (Pitié-Salpêtrière Hospital, Paris, neuropsychologist);

Eye movement: Sophie Rivaud-Pechoux, PhD (ICM, Paris, coordination); Elodie Hainque, MD, PhD (Pitié-Salpêtrière Hospital, Paris);

Sleep assessment: Isabelle Arnulf, MD, PhD (Pitié-Salpêtrière Hospital, Paris, coordination), Smaranda Leu Semenescu, MD (Pitié-Salpêtrière Hospital, Paris), Pauline Dodet, MD (Pitié-Salpêtrière Hospital, Paris);

Genetic data: Jean-Christophe Corvol, MD, PhD (Pitié-Salpêtrière Hospital, Paris, coordination), Graziella Mangone, MD, PhD (Pitié-Salpêtrière Hospital, Paris, co-coordination), Samir Bekadar, MS (Pitié-Salpêtrière Hospital, Paris, biostatistician), Alexis Brice, MD (ICM, Pitié-Salpêtrière Hospital, Paris), Suzanne Lesage, PhD (INSERM, ICM, Paris, genetic analyses);

Metabolomics: Fanny Mochel, MD, PhD (Pitié-Salpêtrière Hospital, Paris, coordination), Farid Ichou, PhD (ICAN, Pitié-Salpêtrière Hospital, Paris), Vincent Perlbarg, PhD, Pierre and Marie Curie University), Benoit Colsch, PhD (CEA, Saclay), Arthur Tenenhaus, PhD (Supelec, Gif-sur-Yvette, data integration);

Brain MRI data: Stéphane Lehericy, MD, PhD (Pitié-Salpêtrière Hospital, Paris, coordination), Rahul Gaurav, MS, (Pitié-Salpêtrière Hospital, Paris, data analysis), Nadya Pyatigorskaya, MD, PhD, (Pitié-Salpêtrière Hospital, Paris, data analysis); Lydia Yahia-Cherif, PhD (ICM, Paris, Biostatistics), Romain Valabregue, PhD (ICM, Paris, data analysis), Cécile Galléa, PhD (ICM, Paris);

DaTscan imaging data: Marie-Odile Habert, MCU-PH (Pitié-Salpêtrière Hospital, Paris, coordination);

Voice recording: Dijana Petrovska, PhD (Telecom Sud Paris, Evry, coordination), Laetitia Jeancolas, MS (Telecom Sud Paris, Evry);

Study management: Vanessa Brochard (Pitié-Salpêtrière Hospital, Paris, coordination), Alizé Chalançon (Pitié-Salpêtrière Hospital, Paris, Project manager), Carole Dongmo-Kenfack (Pitié-Salpêtrière Hospital, Paris, clinical research assistant); Christelle Laganot (Pitié-Salpêtrière Hospital, Paris, clinical research assistant), Valentine Maheo (Pitié-Salpêtrière Hospital, Paris, clinical research assistant).

