## Supplemental material for "Glymphatic defect in isolated REM sleep behavior disorder is associated with phenoconversion to Parkinson’s disease"

**Table S1. Models of the association between ALPS index and trajectories in iRBD**

| **Model Term** | **Conversion to PD^a^** | | | | **Conversion to DLB^a^** | | |
| --- | --- | --- | --- | --- | --- | --- | --- |
|  | **Coefficient (SE)** | **P-value** | | **Odds ratio (95% CI)** | **Coefficient (SE)** | **P-value** | **Odds ratio (95% CI)** |
| *Model 1: ALPS* | | | | | |  |  |
| DTI-ALPS index, left | -0.43 (0.19) | | **0.026** | 0.65 (0.45-0.95) | 0.12 (0.25) | 0.65 | 1.12 (0.68-1.84) |
| Age | -0.001 (0.03) | | 0.98 | 1.00 (0.95-1.05) | 0.03 (0.04) | 0.37 | 1.03 (0.96-1.11) |
| Sex | -0.37 (0.66) | | 0.57 | 0.69 (0.19-2.50) | 0.41 (0.69) | 0.55 | 1.51 (0.39-5.80) |
| Constant | -1.45 (1.75) | | 0.41 | 0.24 | -4.75 (2.53) | 0.06 | 0.01 |
| *Model 2: MoCA* | | | | | | | |
| DTI-ALPS index, left | -0.39 (0.19) | | **0.039** | 0.68 (0.46-0.98) | 0.12 (0.27) | 0.66 | 1.13 (0.66-1.93) |
| Age | 0.01 (0.03) | | 0.78 | 1.01 (0.96-1.06) | 0.01 (0.04) | 0.79 | 1.01 (0.93-1.10) |
| Sex | -0.27 (0.66) | | 0.68 | 0.76 (0.20-2.79) | 0.52 (0.74) | 0.49 | 1.68 (0.39-7.22) |
| MoCA | 0.04 (0.07) | | 0.54 | 1.04 (0.92-1.18) | -0.28 (0.08) | **0.001** | 0.76 (0.65-0.89) |
| Constant | -2.99 (2.64) | | 0.26 | 0.05 | 3.65 (3.63) | 0.31 | 38.41 |
| *Model 3: MDS-UPDRS III* | | | | | | | |
| DTI-ALPS index, left | -0.40 (0.19) | | **0.036** | 0.67 (0.46-0.97) | 0.05 (0.26) | 0.85 | 1.05 (0.63-1.75) |
| Age | -0.01 (0.03) | | 0.79 | 0.99 (0.94-1.05) | 0.03 (0.04) | 0.50 | 1.03 (0.95-1.11) |
| Sex | -0.30 (0.66) | | 0.65 | 0.74 (0.20-2.71) | 0.50 (0.71) | 0.48 | 1.64 (0.41-6.61) |
| MDS-UPDRS III | 0.003 (0.03) | | 0.93 | 1.00 (0.94-1.07) | 0.05 (0.04) | 0.17 | 1.06 (0.98-1.14) |
| Constant | -0.94 (1.78) | | 0.60 | 0.39 | -4.72 (2.72) | 0.08 | 0.01 |

^a^ Tested against any other trajectories in iRBD followed longitudinally

ALPS = glymphatic (‘along the perivascular space’) index; CI = confidence interval; DLB = dementia with Lewy bodies; DTI = diffusion tensor imaging; iRBD = isolated rapid eye movement sleep behavior disorder; MDS = Movement Disorder Society; MoCA = Montreal Cognitive Assessment; PD = Parkinson’s disease; SE = standard error; UPDRS-III = motor part of the Unified Parkinson’s Disease Rating Scale.

**Table S2. Cox proportional hazards model results for predictors of phenoconversion**

| **Variable** | **Coefficient** | **SE** | **HR** | **Lower 95% CI** | **Upper 95% CI** | **P-value** | |
| --- | --- | --- | --- | --- | --- | --- | --- |
| DTI-ALPS index, left | 0.66 | 0.31 | 1.93 | 1.05 | 3.57 | | **0.035** |
| Age | 0.002 | 0.02 | 1.00 | 0.99 | 1.01 | | 0.91 |
| Sex | 0.24 | 0.45 | 1.28 | 0.52 | 3.12 | | 0.59 |

ALPS = glymphatic (‘along the perivascular space’) index; CI = confidence interval; DTI= diffusion tensor imaging; HR = hazard ratio; SE = standard error

**Table S3. Cox proportional hazards model results for predictors of PD conversion**

| **Variable** | **Coefficient** | **SE** | **HR** | **Lower 95% CI** | **Upper 95% CI** | **P-value** |
| --- | --- | --- | --- | --- | --- | --- |
| DTI-ALPS index, left | 0.89 | 0.39 | 2.43 | 1.13 | 5.25 | **0.023** |
| Age | -0.004 | 0.03 | 1.00 | 1.13 | 5.25 | 0.86 |
| Sex | -0.40 | 0.74 | 0.67 | 0.16 | 2.87 | 0.59 |

ALPS = glymphatic (‘along the perivascular space’) index; CI = confidence interval; DTI= diffusion tensor imaging; HR = hazard ratio; SE = standard error

**Table S4. Cox proportional hazards model results for predictors of DLB conversion**

| Variable | Coefficient | SE | HR | Lower 95% CI | Upper 95% CI | P-value |
| --- | --- | --- | --- | --- | --- | --- |
| DTI-ALPS index, left | 0.13 | 0.63 | 1.44 | 0.33 | 3.94 | 0.83 |
| Age | 0.05 | 0.05 | 1.05 | 0.96 | 1.15 | 0.29 |
| Sex | 0.78 | 0.69 | 2.17 | 0.57 | 8.35 | 0.26 |

ALPS = glymphatic (‘along the perivascular space’) index; CI = confidence interval; DTI= diffusion tensor imaging; HR = hazard ratio; SE = standard error

**Figure S1. Kaplan-Meier survival curves comparing low and high glymphatic index for predicting DLB conversion risk**

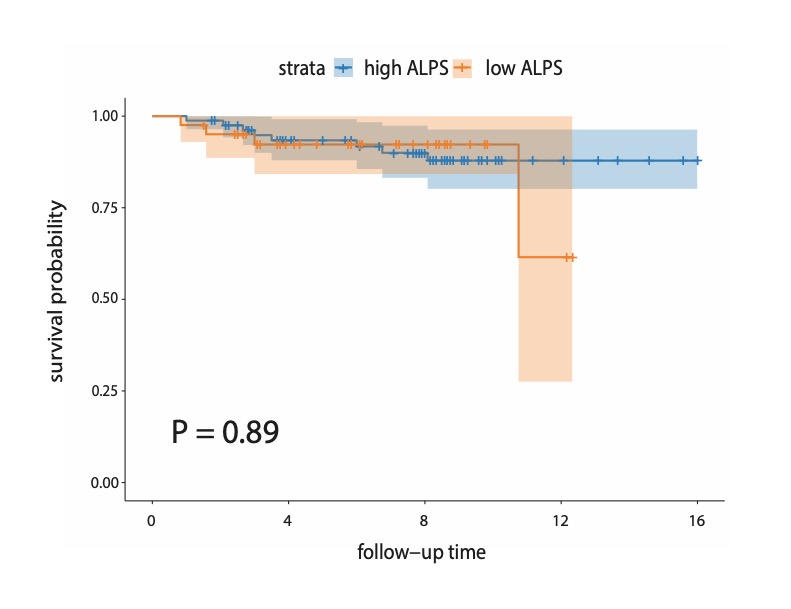

Kaplan-Meier survival curves illustrating the relationship between Low and High ALPS Z-scores and phenoconversion to DLB. A lower left ALPS Z-scores are associated with a higher risk of conversion to PD, with shaded areas representing the 95% confidence intervals.

ALPS = glymphatic (‘along the perivascular space’) index
